# The SENSOR System: Using Standardized Data Entry and Dashboards for Review of Scientific Studies on the Utility of Blood-Based Protein Biomarkers for Patients with Mild Brain Injury

**DOI:** 10.1101/2023.01.10.23284296

**Authors:** Salena Aggerwal, Toufik Safi, Lt (N) Peter Beliveau, Gaurav Gupta

## Abstract

**Background:** There is no objective way of diagnosing or prognosticating acute traumatic brain injuries (TBIs). A systematic review conducted by *Mondello et al*. reviewed studies looking at blood based protein biomarkers in the context of acute mild traumatic brain injuries and correlation to results of computed tomography scanning. This paper provides a summary of this same literature using the SENSOR system.

**Methods:** An existing review written by *Mondello et al*. was selected to apply the previously described SENSOR system (Kamal et al.) that uses a systematic process made up of a Google Form for data intake, Google Drive for article access, and Google Sheets for the creation of the dashboard. The dashboard consisted of a map, bubble graphs, multiple score charts, and a pivot table to facilitate the presentation of data.

**Results:** A total of 29 entries were inputted by two team members. Sensitivities, specificities, positive predictive values (PPVs), negative predictive values (NPVs), demographics, cut-off levels, biomarker levels, and assay ranges were analyzed and presented in this study. S100B and GFAP biomarkers may provide good clinical utility, whereas UCH-L1, C-Tau, and NSE do not.

**Discussion:** This study determined the feasibility and reliability of multiple biomarkers (S100B, UCH-L1, GFAP, C-tau, and NSE) in predicting traumatic brain lesions on CT scans, in mTBI patients, using the SENSOR system. Many potential limitations exist for the existing literature including controlling for known confounders for mild traumatic brain injuries.

**Conclusion:** The SENSOR system is an adaptable, dynamic, and graphical display of scientific studies that has many benefits, which may still require further validation. Certain protein biomarkers may be helpful in deciding which patients with mTBIs require CT scans, but impact on prognosis is still not clear based on the available literature.

## Background

The most common neurological disorders worldwide are traumatic brain injuries (TBIs), and globally their incidence continues to rise (Mondello et al., 2021). In the United States, TBIs cause approximately 370,000 people to be hospitalized, 80,000 annual cases of residual neurological disabilities, and 52,000 deaths (Cervellin et al., 2012).

Despite the high incidence rate of TBIs, efficient and effective in-hospital work up that facilitates decision making and prognosis remains challenging. Computed tomography (CT) is the standard method in the emergency room for diagnosing structural lesions that may require monitoring or surgical interventions in the cases of TBIs. However, CT scans lack sensitivity for mild traumatic brain injury and using the current guidelines only shows clinically relevant lesions in 10% of cases (Asadollahi et al., 2016; Kulbe & Geddes, 2016). CTs scans are also costly and expose patients to radiation which could have long term health effects.

Multiple questions arise when trying to diagnose and treat patients with mild traumatic brain injuries. This includes understaining clinical severity, objective measures to rule emergent consequences, prognostication regarding long term sequelae and how/when to return patients to activities. Given the current tools and technologies this is often challenging and complicated by multiple confounders (e.g. mental health issues, chronicity, number, and/or mechanism of injury). Diagnosis regarding mild traumatic injuries (mTBIs) are often made using subjective criteria like self-reported symptoms (Miller et al., 2022). Investigators have proposed analyzing blood-protein biomarkers in this context to aid with diagnosis as their proposed advantages include better efficiency, portability and cost (Asadollahi et al., 2016).

The five biomarkers primarily studied in the literature are: S100 calcium binding protein B (S100B), glial fibrillary acidic protein (GFAP), neuron specific enolase (NSE), ubiquitin C-terminal hydrolase-L1 (UCH-L1) and Tau.

- S100B is “a low-molecular-weight (9–13 kDa), Ca2+-binding protein primarily found in astrocytic glial cells of the CNS” (Goyal et al., 2013).
- GFAP is “an intermediate filament protein associated with the astroglial cytoskeleton” (Kulbe & Geddes, 2016).
- NSE can also be known as enolase 2 or γ-enolase and is an isomer of the glycolytic enzyme enolases that is present in neuronal cells and cells with neuroendocrine differentiation (*Neuron Specific Enolase - an Overview* | *ScienceDirect Topics*, n.d.).
- UCH-LI is a “biomarker that is highly expressed in the neuronal cytoplasm and is also present in the peripheral nervous system at the neuromuscular junctions” (Kim et al., 2018).
- Tau is a “microtubule-associated protein abundant in axons” that “can be proteolytically cleaved by caspases and calpains to generate a 17kDa fragment [cleaved tau] (c-tau)” (Kulbe & Geddes, 2016).

All of these biomarkers have promising clinical validity and could help in determining the need for CT scans in acute traumatic brain injury (Mondello et al., 2021). Depending on the study, these biomarkers were also used to determine if they could aid in the prognosis, monitoring, and diagnosis of TBIs.

A previous systematic review was conducted by *Mondello et al*. on the clinical application of blood-based protein biomarkers for managing TBIs. These authors consolidated the existing work on how biomarkers could impact the decision around CT scanning in the emergency room. This was done to identify biomarkers that could help in identifying which patients to target with CT scans in order to improve the yield of clinically relevant lesions, monitoring or surgical interventions. The ultimate goal of this work is to improve the mTBI specific assessments by developing methods that may reduce costs, radiation exposure, and enhance the accuracy with which a patient can be diagnosed.

Our study uses the SENSOR system, a process of converting scientific reviews on a particular topic into a dynamic dashboard. A previous study looking at the clinical application of various psychedelics showed the utility of using the SENSOR system in comparison to traditional scientific reviews. The proposed advantages of this system includes improved data organization, and presentation of data in a visual system, which may allow better efficiency of learning and comprehension (Kamal et al., 2022).

## Methods

The articles from the existing systematic review written by *Mondello et al*. were analyzed using the SENSOR system. Their study consisted of 27 articles that analyzed blood-based protein biomarkers in patients with mTBIs (Mondello et al., 2021). While other applications were considered, the main focus was the sensitivity and specificity of the biomarkers in predicting abnormalities on CT scan imaging.

For this paper, two authors (SA and TS) were assigned their own articles, and any concerns regarding data input were discussed with the senior member of the team (GG). A discussion of the review and the cited papers preceded the development of the shared links for the Google form to collect data, Google Drive to access the articles, and Google Sheets to track the intake data (Kamal et al., 2022). Each paper was reviewed once, but 10 articles were randomly selected to be re-reviewed by the other author (SA or TS). This was an additional quality control exercise to ensure that inputs were correct.

One Google Form was made to incorporate all relevant questions used to organize the results of the reviewed studies. Appendix A shows which variables were assigned to short answers, long answers, checkboxes, or multiple choice questions. Questions were structured to facilitate data entry. For simplicity, patients were categorized into the experimental group if their scans were CT positive and the control group if they were CT negative. For numerical inputs, care was taken to standardize input (e.g. sensitivities, specificities, positive predictive values, etc.). Lastly, the form was filled out separate times for articles with multiple biomarkers with relevant information specific to that biomarker only.

All of these articles were made available in a shared Google Drive, and the links to each article were included in the form. The dashboard was created in Google Sheets with filters for the publication year, location (i.e. city), author, control groups, injuries etiology, and the type(s) of biomarkers, and lesions. Additional filters were also included to modify the biomarker’s statistical central tendency measure in the experimental and/or control groups. Score Charts were included to show the following variables; the median cutoff, the assay range, the average length of the trial, the total patients in the trial, the total participants, average age, and gender, between the experimental group and control group. Bubble graphs were also included within the dashboard to compare the sensitivities and specificities of biomarker levels between the experimental group (CT+) and the control group (CT-). Finally, a map and pivot table were included to aid with additional data presentation.

## Results

The data and research examined were extracted from the *Mondello et al*. (2021) meta-analysis on utilization of blood-based protein biomarkers in management of mTBI (Mondello et al., 2021). A total of 27 research paper entries were made by 2 team members (29 entries in total). The patients included in these studies were assessed in the emergency department post injury, and for whom blood samples were collected, and analyzed for any of the six following protein biomarkers: S100B, UCH-L1, GFAP, C-tau, neurofilament proteins, and NSE.

Appendix A displays the variables included in the Google Form for these 27 studies, prior to creation of the SENSOR System dashboard. More data was inputted into the forms than is displayed in the dashboard. See Figure 1 for a global view of the dashboard. The dashboard allows the data to be displayed in various ways using the slicers.

**Figure 1:**
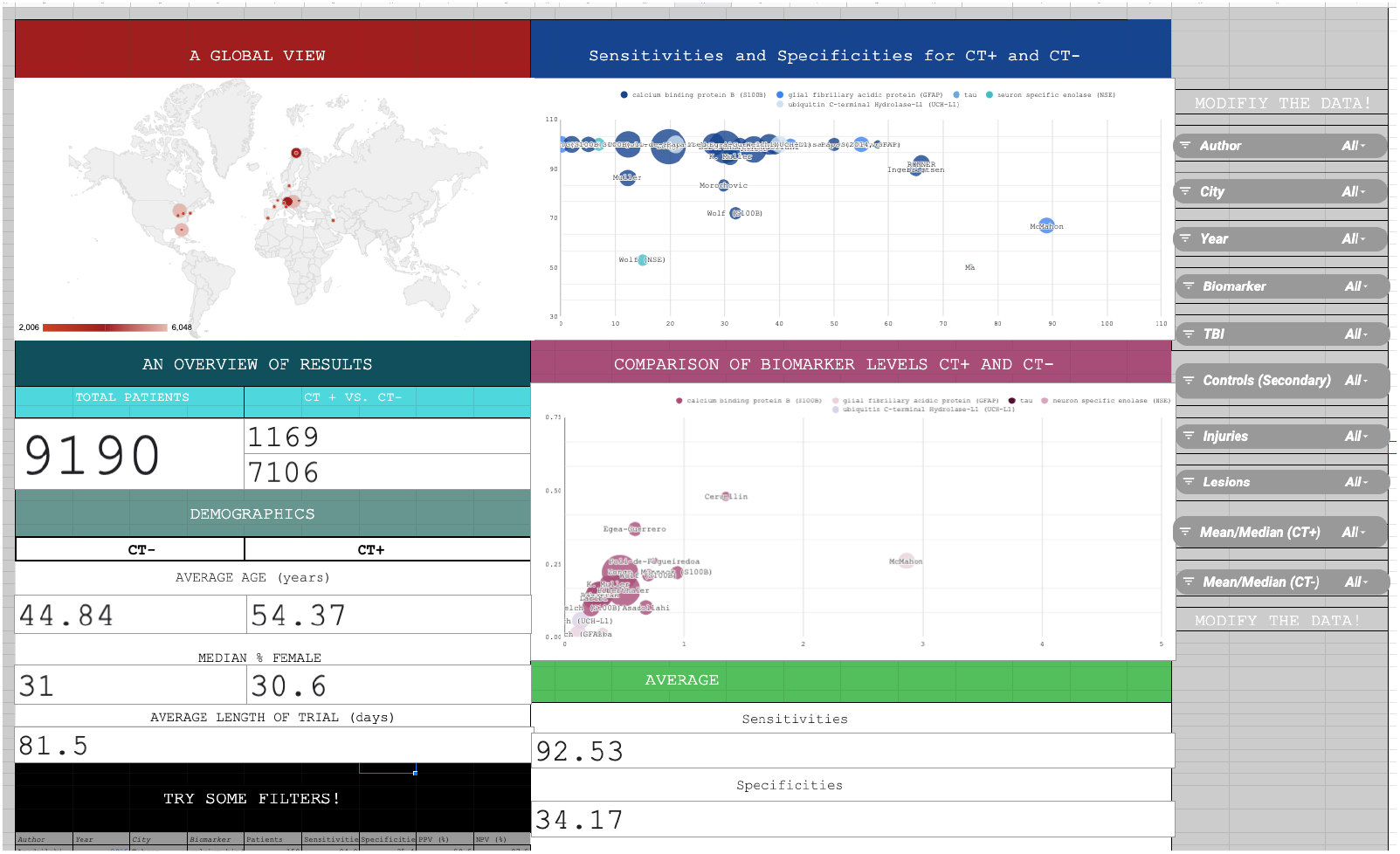
Global View of Dashboard. Link to dashboard: https://docs.google.com/spreadsheets/d/1Zar8WqPNdPhNdyPfi1AswNVaMwCGBNfC739IJy4_E2o/edit?usp=sharing

**Table 1:** Results in the SENSOR System for the Blood-Based Protein Biomarkers (ie. S100B, GFAP, UCH-L1, C-Tau, and NSE)

| Biomarker | Sensitivity Range (%) | Specificity Range (%) | Number Input |
| --- | --- | --- | --- |
| S100B | 83.3-100 (*majority above 90) | 0-55 | 20 |
| GFAP | 67-100 | 0-89 | 4 |
| UCH-L1 | 100 | 21-40 | 2 |
| C-Tau | 50 | 75 | 1 |
| NSE | 53-100 | 6.9-15 | 2 |

S100B is the most researched biomarker, in contrast to the other markers (Kulbe & Geddes, 2016). The data is consistent with S100B having high sensitivity for mTBI. GFAP shows good prognostic value for mTBIs and could potentially be a more sensitive marker than S100B when identifying intracranial lesions (Kulbe & Geddes, 2016). Other biomarkers appear to have poor prognostic biomarkers for mTBIs (Kulbe & Geddes, 2016). Few studies looked at S100B and C-Tau being a possible predictor of post-concussion syndrome (PCS). The results concluded that elevated S100B and C-Tau levels have no significant correlation with the prognosis of PCS for mTBIs. Further research and additional trials is necessary to fully determine the clinical application of these biomarkers.

## Discussion

The main goal of this study was to apply the SENSOR system to determine the feasibility and reliability of multiple biomarkers (S100B, UCH-L1, GFAP, C-tau, and NSE) in predicting lesions in acute traumatic brain lesions on CT scans.

The study specifically examined the technical process of data input and dashboard using an existing review article. As previously noted, several scientific and clinical domains could benefit from applying the SENSOR system as an alternative to the conventional dissemination of scientific reviews.

The applications might incorporate conventional review articles, journal or organization-specific dashboards summarizing current literature. It could then be made available to certain authors exclusively, or systems created to guide future research for validation and discovery. This provides a chance to standardize reporting, concentrate legacy datasets, speed the submission process, promote researcher collaboration, quantify relative contribution of participating authors, and improve patient involvement as the technology and protocols for these systems develop.

To manage these systems over time, organizations must ensure that the proper infrastructure is in place. Understanding the resources and effort required for the various components, such as data entry, dashboard maintenance, and dissemination, is necessary for this (Kamal et al., 2022). Further investigation would be necessary to properly test usability, and applicability, when compared to conventional review articles.

The main review article examined, through the sensitivity and specificity of various biomarkers graphically. This dashboard shows that only S100B, out of the five biomarkers investigated, assists in making well-informed decisions for patients presenting to the emergency department (ED) with suspected cerebral lesions following mTBI (Mondello et al., 2021).

With further research, S100B and GFAP could potentially be used for clinical application. C-Tau, UCH-L1, and NSE have shown to have poor prognosis value. Very few studies examined C-tau, UCH-L1, and NSE and further research would need to be conducted before making conclusive statements.

Limitations of the reviewed articles include variation in study design, time of admittance to the ED, number of biomarkers analyzed, secondary analyses if present, and outcome measures, which affects the accuracy of the data provided. In addition, there were concerns regarding specificity in S100B, GFAP, and NSE, along with contrasting data regarding using UCH-L1 and Tau as a biomarker of mTBI (Kim et al., 2018; Kulbe & Geddes, 2016). There was also variation in exclusion and inclusion criteria in the trials (i.e. one or more of the papers did not exclude head injuries with skull fractures and included severe and/or moderate head injury).

As noted, the review by *Mondello et al*. primarily considered studies on biomarkers identifying structurally relevant CT scan lesions for acute management of complicated mTBI. However, identifying objective tools for diagnosing and prognosticating around mTBI is far more complex. This includes looking at existing work on biomarker applications outside of imaging and other objective measures and their implications for the prognosis, monitoring, and diagnosis of TBIs in a wide range of populations (e.g. civilian, military, and athlete populations). Moreover, age, sex, biological cofounders, inflammation, and extracranial lesions could potentially have implications for pathophysiology and therefore could amplify the importance of these biomarkers for mTBIs (A. P. Di Battista et al., 2020; A. P. Di Battista, Moes, et al., 2018; Kim et al., 2018; Kulbe & Geddes, 2016; Pyndiura et al., 2020).

Military personnel and athletes are more susceptible to TBIs (Kulbe & Geddes, 2016). The biomarkers discussed in this paper (ie. UCH-L1 and Tau) could potentially aid military occupational medicine by being a helpful assessment tool for low-level over-pressure exposure and concussion-like breacher’s neuropsychiatric symptoms (Boutté et al., 2021). In the military population, other biomarkers such as metabolites (ie, acetic acid, formate, creatine, acetone, methanol, and glutamic acid) could also aid blast injury diagnosis, surveillance, and avoidance (Miller et al., 2022).

Identification of these biomarkers could benefit athletes by (a) helping to characterize pathophysiology and secondary injury after sport-related concussions (SRC) and (b) clinical application for early detection of concussions (Battista et al., 2018; A. P. Di Battista, Rhind, et al., 2018). Both groups could benefit from these biomarkers being used to detect the neurological injury associated with repetitive subconcussive head trauma (Oliver et al., 2018)..

In athletes, other biomarkers such as brain derived neurotrophic factor (BDNF), choline, dehydroepiandrosterone sulfate (DHEA-S) interleukin IL-6, monocyte chemoattractant protein (MCP)-1 and (MCP)-4, myo-inositol, N-acetyl aspartate (NAA), peroxiredoxin-6 (PRDX-6), serum NF-L, and Willebrand factor (vWF), could be used to prognosis SRC and/or repetitive concussions (Battista et al., 2016, 2018, 2020; Churchill et al., 2017; A. P. Di Battista, Rhind, et al., 2018; A. P. Di Battista et al., 2019; Oliver et al., 2018).

Biomarkers could potentially be used to aid in prognosis in civilian populations as well. Studies thus far have looked at various biomarkers including; brain derived neurotrophic factor (BDNF), catecholamine, creatine kinase-BB isoenzyme (CKBB), monocyte chemoattractant protein (MCP)-1, myelin based proteins (MBP), matrix metalloproteinase (MMP)-9, Neutrophil gelatinase-associated lipocalin (NGAL), neurogranin (NRGN), peroxiredoxin 6 (PRDX6), SBDP145, TNFα, VEGF, visinin-like protein (VILIP)-1, von Willebrand factor (vWF) (Buonora et al., 2015; A. Di Battista et al., 2013; A. P. Di Battista et al., 2015; A. P. Di Battista, Moes, et al., 2018; Edwards et al., 2020; Kim et al., 2018; Rizoli et al., 2017).

Future research could also focus on both physiological and psychological associations. This is important because previous research has shown (a) an association of concussion history with an increased risk of clinically diagnosed depression and depressive symptoms later in life in former athletes and (b) possible comorbidity between blast-induced TBIs and PTSD and/or depression (Hutchison et al., 2018; Vartanian et al., 2020).

## Conclusion

The SENSOR system can be used to substitute and/or supplement the dissemination of summaries through conventional scientific reviews. The adaptable, dynamic, and graphical display has many claimed benefits, all of which need further validation.

All five biomarkers analyzed in this study would need further research before widespread clinical application. S100B is the most well-researched biomarker and has been seen to be reliable in predicting traumatic intracranial lesions.

Therefore, it could potentially be used to make informed decisions like when to conduct CT scans on acute mTBI patients. GFAP is not as well researched as S100B, but with further research, it could potentially be a more sensitive marker than S100B when identifying intracranial lesions. UCH-L1, C-Tau, and NSE have shown poor prognosis values. More trials are necessary to validate the reliability of these findings.

## Data Availability

All data produced in the present work are contained in the manuscript

### Appendix A: Google Form Variables and Questions Subtype Assignment

| Short Answer | Long Answer | Checkboxes | Multiple Choice |
| --- | --- | --- | --- |
| Email<br>Date<br>Year of Publication<br>Author (Last name)<br>G-Drive article link<br>Primary City<br>Country(ies)<br>Median age of all participants<br>Median age of control group<br>Median age of experimental group<br>Gender (% female) for control group<br>Gender (% female) for experimental group<br>Gender (% female) for all participants<br>Total Number of Patients in trial<br>Total Number of participants in experimental group<br>Total Number of participants in control group<br>Type of TBI (primary)<br>Type of TBI (secondary)<br>Total length of trial<br>Number of days participants participated<br>Prevalence of CT scan findings (%)<br>Number of times sample was collected<br>Max time post injury<br>Mean/Median Biomarker levels in patients with CT positive (ug/L)<br>Mean/Median Biomarker levels in patients with CT negative (ug/L)<br>positive predictive value (%) for CT +/-<br>negative predictive value (%) for CT +/-<br>Sensitivities (%) for secondary measure<br>Specificities (%) for CT +/-<br>Specificities (%) for secondary measure<br>Assay Range (ug/L)<br>Cutoff (ug/L) | Inclusion Criteria<br>Exclusion Criteria<br>Comments | Number of Biomarkers<br>Type of Biomarker<br>Context of people injured<br>Nature of controls for primary dashboard<br>Nature of controls for secondary dashboard<br>Type of lesions shown on CT+ scans<br>Limitations | How the samples were processed<br>Biomarker Level Central Tendency Measure for experimental group<br>Biomarker Level Central Tendency Measure for control group |

